# Validation of a Novel Molecular Assay to the Diagnostic of COVID-19 Based on Real Time PCR with High Resolution Melting

**DOI:** 10.1101/2021.07.14.21260471

**Authors:** Beatriz Iandra da Silva Ferreira, Natália Lins da Silva Gomes, Wagner Luis da Costa Nunes Pimentel Coelho, Vanessa Duarte da Costa, Vanessa Cristine de Souza Carneiro, Rafael Lopes Kader, Marisa Pimentel Amaro, Lívia Melo Villar, Fábio Miyajima, Soniza Vieira Alves-Leon, Vanessa Salete de Paula, Luciane Almeida Amado Leon, Otacilio Cruz Moreira

**Author notes:** Corresponding author Otacilio Cruz Moreira. Real Time PCR Platform RPT09A, Laboratory of Molecular Biology and Endemic Diseases, Oswaldo Cruz Institute/ Fiocruz, Rio de Janeiro, Brazil. Funding: This work has been founded by INOVA Fiocruz, Coordenação de Aperfeiçoamento de Pessoal de Nível Superior (CAPES - Finance Code 001), CNPq and FAPERJ. LMV, SVAL, VSP, OCM are research Fellows from CNPq and FAPERJ (JCNE and CNE). LAAL is a researcher fellow from FAPERJ (JCNE).

## Abstract

With the emergence of the Covid-19 pandemic, the world faced an unprecedented need for RT-qPCR-based molecular diagnostic tests, leading to a lack of kits and inputs, especially in developing countries. Hence, the costs for commercial kits and inputs were overrated, stimulating the development of alternative methods to detect SARS-CoV-2 in clinical specimens. The availability of the complete SARS-CoV-2 genome at the beginning of the pandemic facilitated the development of specific primers and standardized laboratory protocols for Covid-19 molecular diagnostic. High-sensitive and cost-effective molecular biology technique based on the Melting Temperature differences between purine and pyrimidine bases can be used to the detection and genotyping of pathogens in clinical specimens. Here, a RT-qPCR assays with High Resolution Melting (HRM-RTqPCR) was developed for different regions of the SARS-CoV-2 genome (RdRp, E and N) and an internal control (human RNAse P gene). The assays were validated using synthetic sequences from the viral genome and clinical specimens (nasopharyngeal swabs, serum and saliva) of sixty-five patients with severe or moderate COVID-19 from different states in Brazil, in comparison to a commercial TaqMan RT-qPCR assay, as gold standard. The sensitivity of the HRM-RTqPCR assays targeting N, RdRp and E were 94.12, 98.04 and 92.16%, with 100% specificity to the 3 targets, and diagnostic accuracy of 95.38, 98.46 and 93.85%, respectively. Thus, the HRM-RTqPCR emerges as an alternative and low-cost methodology to increase the molecular diagnostic of patients suspicious for Covid-19, especially in restricted-budget laboratories.

## INTRODUCTION

Severe acute respiratory syndrome coronavirus 2 (SARS-CoV-2) is a positive-sense single-stranded RNA virus ^1^ causative of coronavirus disease 2019 (COVID-19), the airborne-transmitted respiratory illness responsible for the COVID-19 pandemic ^2^. Since its emergence in December 2019, until July 5^th^ 2021, WHO has reported over 183 million cases and about 4 million deaths from COVID-19 ^3^. Compared to the last WHO weekly report, the highest numbers of new cases were reported from India, Brazil, the United States of America, Turkey and Argentina. The majority (∼ 80%) of patients have mild symptoms or are asymptomatic, and about 20% of cases may require hospital care because they have difficulty breathing. The mortality rate of COVID-19 has been estimated at around 3% ^4^.

The availability of the complete SARS-CoV-2 genome at the beginning of the pandemic facilitated the development of specific primers and standardized laboratory protocols for COVID-19 molecular diagnostic ^5,6^. The protocols for the firsts real-time RT-qPCR assays, targeting the SARS-CoV-2 RNA-dependent RNA polymerase (RdRp), envelope (E) and nucleocapsid (N), were published on January, 2020 ^7^. After that, WHO produced a technical manual ^8^, where seven in house RT-PCR assays are recommended, developed by CDC-China, Pasteur Institute-France, CDC-USA, NIID-Japan, Charité-Germany, HKU-Hong Kong and NIH-Thailand. All the recommended protocols using TaqMan systems, with TaqMan probes designed for the different regions of the viral genome. Of these, one assay has an internal human control (RNAse P gene, CDC-USA protocol), capable of monitoring the quality of the nucleic acid extraction and validating the true negative result. Based on these reactions, a variety of molecular diagnostic kits were produced and validated in record time. Even so, the production of these kits could be not enough to cover the immense demand for diagnostics worldwide, and the costs are generally overpriced. Therefore, it should be considered to invest in low-cost alternative technologies, to assist the diagnosis of COVID-19 in a less dependent way on imported products and inputs.

The reverse transcription polymerase chain reaction real time assays with High Resolution Melting (HRM-RTqPCR) analysis is a high-sensitive and cost-effective molecular biology technique based on the Melting Temperatures (Tm) difference between purine and pyrimidine bases that can be used for the detection of mutations, polymorphisms and epigenetic differences (quantification of methylation status of CpG sites) in double-stranded DNA samples ^9^. HRM-qPCR based methodologies have been reported for the detection and genotyping of different pathogens, such as virus, bacteria and protozoa in clinical samples ^10-13^. Due to the possibility of PCR products differentiation by the Tm at the melting curves with high resolution, it is possible to combine different sets of primers, specific for different pathogens or more than one target in the same pathogen and/or an internal control. HRM-RTqPCR master mix contains a fluorophore instead of fluorescent probes, for this reason it becomes less expensive than TaqMan assays or DNA sequencing to the molecular diagnostic or pathogens genotyping from clinical samples.

In this study, real-time RT-PCR assays with High Resolution Melting were developed for different regions of the SARS-CoV-2 genome (RdRp, E and N) and an internal control (human RNAse P gene). The assays were standardized and validated using synthetic sequences from the viral genome and clinical specimens (nasopharyngeal swabs, serum and saliva) of patients with clinical suspicion of COVID-19 from different states of Brazil, in comparison to TaqMan RT-qPCR assays, as gold standard. Therefore, we aimed to obtain an sensitive and specific assay to COVID-19 diagnosis as an alternative to TaqMan assay to overcome high costs and the limited supplies that might delay accurate diagnosis.

## MATERIAL AND METHODS

### Ethics Statement

This study was approved by the ethical committee of Clementino Fraga Filho University Hospital, from Federal University of Rio de Janeiro (UFRJ), CAAE number 31240120.0.0000.5257. All patients or a member of their families signed the consent form.

### Clinical samples

In this study, a panel of 42 nasopharyngeal swab, 12 serum and 11 saliva samples from 65 patients with severe or moderate COVID-19, admitted during the acute phase of the infection, were included. Fourty-Five and Twenty patients were from the Rio de Janeiro state and Ceará state, respectively. Of these samples, 51 were positive and 14 were negative to the molecular detection of SARS-CoV-2 for the RT-qPCR TaqMan assay targeting the N region of viral genome. The inclusion criteria was the presence of clinical symptoms compatible with COVID-19 during the admittance and presented at least one SARS-COV-2 laboratorial analysis. Patients under 18 years old, pregnant women and with cancer were excluded from this study.

To the nasopharyngeal samples collection, swab was inserted through the nostril with a rotation movement until the nasopharynx was reached, and the sample was obtained by rotating the swab gently for 2-3 seconds. Then, the swab was placed into a 5 ml tube containing 3 ml viral transport medium (VTM). After transportation to the laboratory, tubes containing the swabs were vortexed vigorously, VTM was transferred to 1.5mL tubes and stored at −80°C until RNA extraction.

To the saliva samples collection, patients were asked to self-collect the saliva in a sterile dry tube, closing the lid after placing the saliva in it. The staff cleaned the outside of the tube with 1/10 diluted bleach-impregnated cloth, after taking the container while wearing gloves. After taking both samples, they were delivered to the laboratory inside the triple transport system.

To the serum samples collection, around five milliliters of blood were harvested in a BD Vacutainer® Plus Plastic Serum tube and remained resting for 30 min at room temperature, to generate the clot from blood cells and the serum phase. After that, tubes were centrifuged at 1,000 x g for 10 minutes at 4 °C and serum was collected and stored at −80°C until use.

### Viral RNA extraction and cDNA synthesis

RNA was extracted from 140 µL of nasopharyngeal swab VTM (Virus Transport Media), serum or saliva, using the QIAamp Viral RNA Mini Kit (Qiagen, USA), according to the manufacturer’s instructions. At the final step of the protocol, RNA was eluted at 60 µL of AVE Buffer and stored at −80°C until use.

To the two-steps HRM-RTqPCR assays, reverse transcription was performed from 5 µL RNA using the SuperScript III First-Strand Synthesis System (Invitrogen, USA), according to manufacturer’s instructions, using 1 µL of the equimolar mixture of 2019-nCoV_N2-R, nCoV 1 _IP4-14146Rv, E_Sarbeco_R2 and RP-R primers at 2 µM each.

### Real Time RT-PCR with TaqMan assays

All patient samples were analyzed for SARS-Cov-2 N2 and human RNAseP targets using a commercial One-Step RTqPCR TaqMan kit, 2019-nCov CDC RUO Kit (IDT DNA, USA. Cat. No. 10006625), according to manufacturer’s instructions. RT-qPCR was carried out in 10 µL reaction containing 2 µL RNA, 5 µL Promega Go-Taq Probe-1-Step-RT-qPCR System [2X], 0.2 µL GoScript RT mix for 1-step RT-qPCR, 0.75 µL Mix Primer-Probe FAM/BHQ (N2 or RNAse P) TaqMan, 0.25 µL CXR Reference Dye and 1.8 µL ultrapure water. Real-time PCRs were carried out on Applied Biosystems Quantstudio 3 Real-Time PCR System (ThermoFisher, USA) using the following cycling conditions: 15 min at 50 °C, 10 min at 95 °C, followed by 40 cycles of 15 sec at 95°C and 60 sec at 55°C. Fluorescence was collected after each cycle at the annealing/extension step. All samples were run in duplicate and threshold was set at 0.02 for analysis.

### Primers selection

To select the primers for HRM-RTqPCR assays (Table 1), sequences were analyzed for Tm, hairpin, self-dimer and hetero-dimer formation using the OligoAnalyzer Tool (Integrated DNA Technologies, USA), available at https://www.idtdna.com/calc/analyzer. In addition, primers were also evaluated *in silico* for their specificity. A primer-BLAST search (https://www.ncbi.nlm.nih.gov/tools/primer-blast/) using the sequences of each primer pair and allowing up to 4 mismatches (none in the 3’
s end) in the complete set of the NCBI database (non-redundant sequences) resulted only in SARS-CoV-2 and RNAse P gene related-sequences.

**Table 1.**
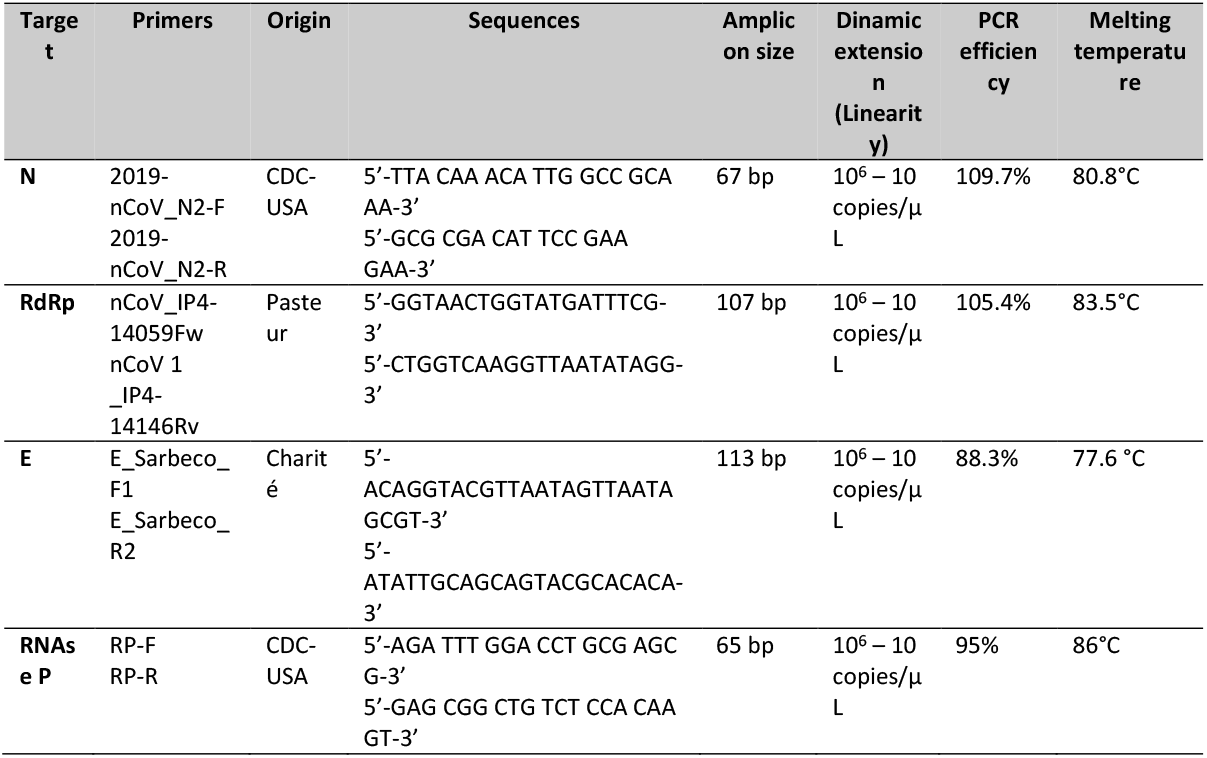
Primers, linearity and melting curves parameters for SARS-CoV-2 and human RNAse P amplification.

### Real Time RT-PCR with High Resolution Melting

To the HRM-RTqPCR assays, all cDNA were analyzed for SARS-CoV-2 N2, RdRp and E target. As an internal control, the human RNAse P target was used. The assays were carried out in a final volume of 10 µL, containing 2 µL [5X] Hot Firepol Evagreen HRM Mix (No ROX), 200 nM 2019-nCoV_N2 F and 2019-nCoV_N2 R primers; or 300 nM nCoV_IP4-14059Fw and nCoV_IP4-14059 Rv primers; or 200 nM E_Sarbeco_F1and 300 nM E_Sarbeco_R2 primers (Table 1); or 300 nM RP-F and 300 nM RP-R primers; 2 µL cDNA and ultrapure water to reach 10 µL. Real-time PCRs were carried out on Applied Biosystems Quantstudio 3 Real-Time PCR System (ThermoFisher, USA) using the following cycling conditions: 10 min at 95 °C, followed by 40 cycles of 15 sec at 95°C and 60 sec at 58 °C (N and RdRp targets) or 60 °C (E and RNAse P targets), where fluorescence was collected after each cycle. Melt curve stage conditions were: 10 sec at 95°C, 60 sec at 60 °C, 15 sec at 95 °C (where fluorescence was collected at 0.025 °C/s rate) and 15 sec at 60 °C. Threshold was stablished and set at 10,000 for analysis.

### Controls and Analysis

In each assay, positive and negative controls were used, as follows: low positive control (synthetic template of single-strand DNA containing N, RdRp and E genes of SARS-CoV-2 (GenBank NC_045512.2), at 200 copies/µL), very low positive control (synthetic template of single-strand DNA containing N, RdRp and E genes of SARS-CoV-2 (GenBank NC_045512.2), at 20 copies/µL), and negative template control. In addition, one negative control of RNA extraction was added at each extraction batch, using ultrapure water instead samples. To observe the amplification plots and Ct values, results were analyzed using the QuantStudio Design and Analysis Software v1.5.1 (Applied Biosystems, USA). High Resolution Melt Software v.3.2 (Applied Biosystems, USA) was used to distinguish the high resolution melting curves and obtain Melting Temperature (Tm) values.

### Analytical Validation

The analytical validation (Dinamic Extension, Precision, Reproducibility) assays were performed using synthetic single strand DNA molecules containing the N, RdRp and E regions of SARS-CoV-2 from the Severe acute respiratory syndrome coronavirus 2 isolate Wuhan-Hu-1 (GenBank NC_045512.2). To the linearity assay, a 1:10 serial dilution of the synthetic DNA was performed from 10^6^ to 1 copies/µL in TE buffer. To the precision assay, three concentrations of the synthetic SARS-CoV-2 templates were used (12, 10 and 8 copies/µL). Forty technical replicates were assayed in each concentration to the same operator in the same day, and the results were compared. To the Reproducibility assay, the same three concentrations of the synthetic sequences, at 12, 10 and 8 copies/µL, were also used. The amplification was performed in 40 technical replicates for each template concentration, divided in two consecutive days, by the same operator.

### Statistical analysis

All experiments were performed at least in two technical replicates. Data distribution was evaluated by the Shapiro-Wilk normality test. Student’s t-test or Mann– Whitney Rank Sum test was used to analyze the statistical significance of the observed differences (according to the parametric or nonparametric distribution of the values, respectively) with SigmaPlot for Windows version 12.0 (Systat Software, Inc). Values of sensitivity, specificity, PPV, NPV, Diagnostic accuracy and Cohen’s kappa coefficient were calculated using the Open Source Epidemiologic Statistics for Public Health (OpenEpi software V3.01), available at www.openepi.com. Results were expressed as means and standard deviations, and differences were considered significant if p < 0.05.

## RESULTS

### Primers selection

To select the appropriate primers to develop the HRM-RTqPCR assays, the primers designed for COVID-19 molecular diagnostic described by WHO ^8^ were evaluated based on the Tm similarity between primer pairs, low hairpin, self-dimer and hetero-dimer formation tendency, small amplicon size (below 120 bp), and PCR efficiency. Therefore, they were selected the primers 2019-nCoV_N2-F and 2019-nCoV_N2-R (CDC, USA), nCoV_IP4-14059Fw and nCoV 1 _IP4-14146Rv (Institut Pasteur, France), E_Sarbeco_F1 and E_Sarbeco_R2 (Charité, Germany) and RP-F and RP-R (CDC, USA) targeting N, RdRp and E regions of the SARS-Cov-2 genome and Human RNAse P gene, respectively (Table 1). Those primers were selected based on the Tm similarity between primer pairs, low hairpin, self-dimer and hetero-dimer formation tendency and small amplicon size (below 120 bp).

### Analytical Validation of HRM-RTqPCR assays

After standardization, HRM-qPCR assays were initially validated using synthetic DNA sequences of SARS-CoV-2 (GenBank NC_045512.2). A single and sharp peak at the derivative HRM curve was generated to each target, with Tm 80.8, 83.5, 77.6 and 86°C to N, RdRp, E and RNAse P, respectively (Table 1 and Fig. 1A). Due to the distribution of the peaks, the pre-melt and post-melt regions of HRM were stablished before 72 and after 90.8 °C, respectively (Fig. 1A). Thus, the normalized HRM graph showed four distinguished curves, capable to be analyzed in conjunct (Fig. 1B). The analysis of the standard curves produced by the serial dilution of the synthetic controls showed a dynamic range between 10^6^ and 10 copies/µL to the N, RdRp and E SARS-CoV-2 targets (Table 1 and Fig. 1C). The PCR efficiencies were between 88.3 and 109.7%, leading to the possibility of viral load quantification by HRM-RTqPCR with accuracy, with low Ct difference between the three targets. In addition, it was possible to observe the same approximate limit of detection (LOD) – the lowest concentration detected in the linearity assay, around 10 copies/ µL, to the three targets in SARS-CoV-2.

**Figure 1.**
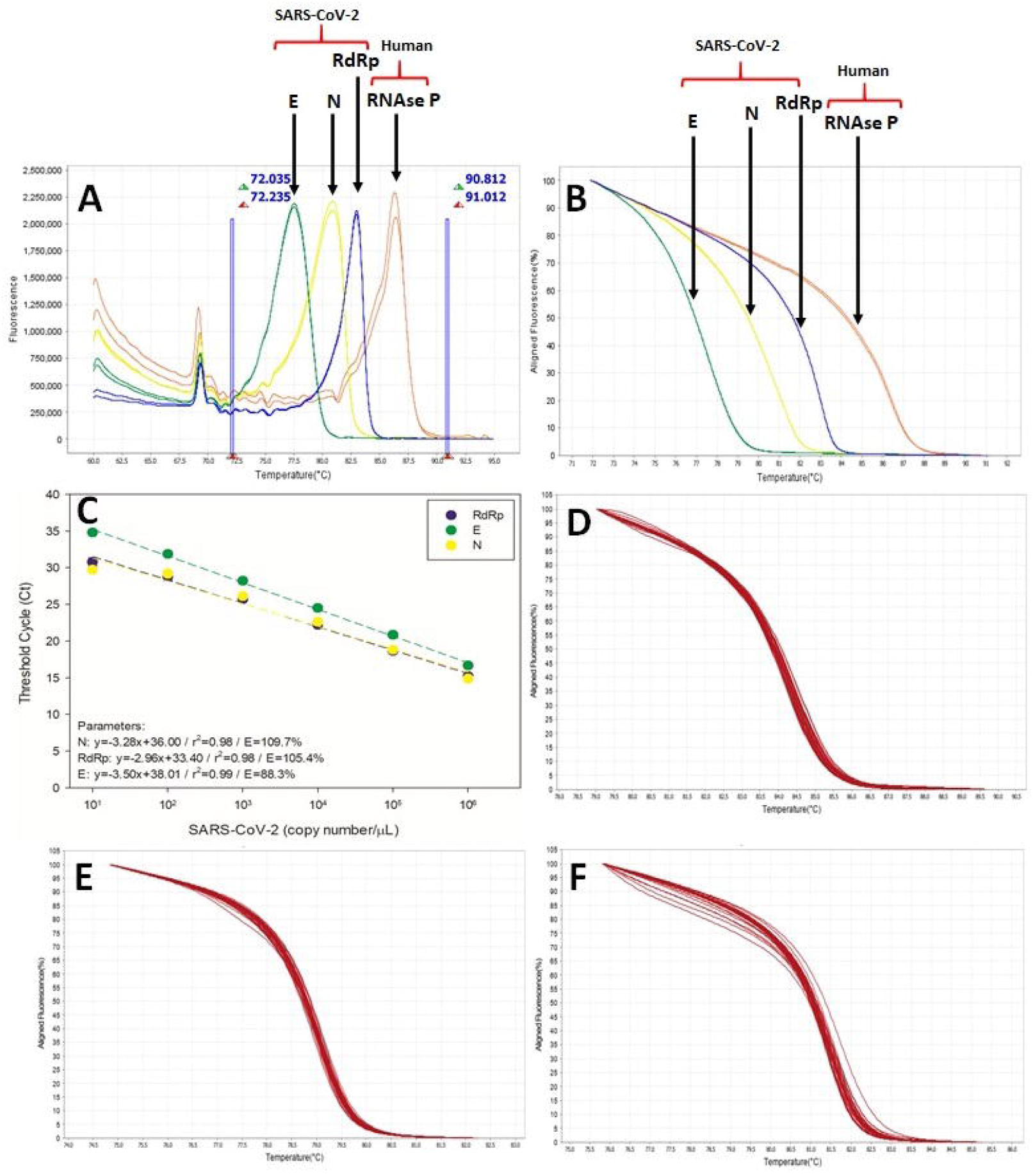
HRM-RTqPCR assays for SARS-CoV-2 E, N and RdRp targets. **A**. Derivative HRM curves for SARS-CoV-2 E (green), N (yellow) and RdRp (blue) and Human RNAse P (red) targets. The vertical blue lines divide the Pre-melt, Melt and Post-melt regions. **B**. Normalized HRM curves for SARS-CoV-2 E (green), N (yellow) and RdRp (blue) and Human RNAse P (red) targets. **C**. Dinamic extension (linearity) for the SARS-CoV-2 E (green), N (yellow) and RdRp (blue) targets amplification. **D**. Normalized HRM curves for SARS-CoV-2 N region from COVID-19 clinical samples. **E**. Normalized HRM curves for SARS-CoV-2 RdRp region from COVID-19 clinical samples. **F**. Normalized HRM curves for SARS-CoV-2 E region from COVID-19 clinical samples.

When clinical samples (RNA extracted from nasopharyngeal swab, serum or saliva) were evaluated, the same normalized HRM curve profile was observed in comparison to the synthetic controls, with few differences between samples (Fig. 1D, 1E and 1F to the N, RdRp and E targets, respectively), validating the use of the HRM curves to confirm the product of amplification of SARS-CoV-2 in the positive samples.

Following analytical validation, the precision of HRM-RTqPCR assays was evaluated. For that, three concentrations around the estimated LOD (10 copies/µL) were selected: 12, 10 and 8 copies/µL of the synthetic SARS-CoV-2 templates. For N, RdRp and E targets, forty technical replicates were assayed in each concentration, and the results were compared (Table 2). To the N target, 100% of positive amplification was observed to the three concentrations, with Ct means (± Standard Deviation) of 29.80±0.51, 30.44±0.43 and 30.57±0.53, and coefficients of variation of 1.73%, 1.40% and 1.74%, respectively. To the RdRp target, 97.5% of positive amplification was observed to the three concentrations, with Ct means of 30.85±0.51, 30.77±0.99 and 30.70±0.83, and coefficients of variation of 3.74%, 3.20% and 2.71%, respectively. Finally, to the E target, 97.5%, 87.5% and 97.5% of positive amplification were observed to 12, 10 and 8 copies/µL, with Ct means of 28.87±0.46, 28.41±0.36 and 28.70±1.05, respectively. To this target, the coefficients of variation were 1.60%, 1.27% and 3.65%, respectively. Even with slight differences, the coefficient of variation to all targets and concentrations tested were lower than 5.0%, showing the high precision of the assays.

**Table 2.** Precision of HRM-RTqPCR assays targeting SARS-CoV-2 E, N and RdRp. SARS-CoV-2 templates were assayed in 40 technical replicates, at 12, 10 and 8 copies/µL.

| Parameter | Sample concentration |  |  |
| --- | --- | --- | --- |
|  | 12 copies/μL | 10 copies/μL | 8 copies/μL |
| **N target** |  |  |  |
| Positive samples, n=40 | 100% | 100% | 100% |
| Ct mean | 29.80 | 30.44 | 30.57 |
| Standard Deviation | 0.5145 | 0.4274 | 0.5302 |
| Coefficient of variation | 1.726% | 1.404% | 1.735% |
| **RdRp target** |  |  |  |
| Positive samples, n=40 | 97.5% | 97.5% | 97.5% |
| Ct mean | 30.85 | 30.77 | 30.70 |
| Standard Deviation | 1.155 | 0.9851 | 0.8307 |
| Coefficient of variation | 3.743% | 3.202% | 2.706% |
| **E target** |  |  |  |
| Positive samples, n=40 | 97.5% | 87.5% | 97.5% |
| Ct mean | 28.87 | 28.41 | 28.70 |
| Standard Deviation | 0.4606 | 0.3599 | 1.046 |
| Coefficient of variation | 1.595% | 1.267% | 3.646% |

The reproducibility of the HRM-RTqPCR assays on the detection of SARS-CoV-2 was also evaluated throw the amplification (Ct values) of the synthetic sequences at 12, 10 and 8 copies/µL. The amplification was performed in 40 technical replicates for each template concentration, divided in two consecutive days, by the same operator. The box plot in Fig. 2 shows the distribution of Ct values in the technical replicates and between the two days of analysis, for each target. To the N target, the medians of Ct values for days one and two, at 12 copies/µL, were respectively 29.57 and 29.89. At 10 copies/µL, the medians were 30.46 and 30.34 and, at 8 copies/µL, the medians were 30.39 and 30.63, respectively. To the RdRp target, the medians of Ct values for days 1 and 2, at 12 copies/µL, were respectively 31.49 and 29.92. At 10 copies/µL, the medians were 30.42 and 30.84 and, at 8 copies/µL, the medians were 30.40 and 30.78, respectively. To the E target, the medians of Ct values for days one and two, at 12 copies/µL, were respectively 28.58 and 28.93. At 10 copies/µL, the medians were 28.25 and 28.62 and, at 8 copies/µL, the medians were 28.42 and 28.61, respectively. It was possible to observe a little dispersion of Ct values in the replicates, and a small number of outliers in all concentrations tested. In addition, no significant difference was observed in Ct values between day 1 and day 2 for all the targets, which means a good reproducibility of the HRM-qPCR assays.

**Figure 2.**
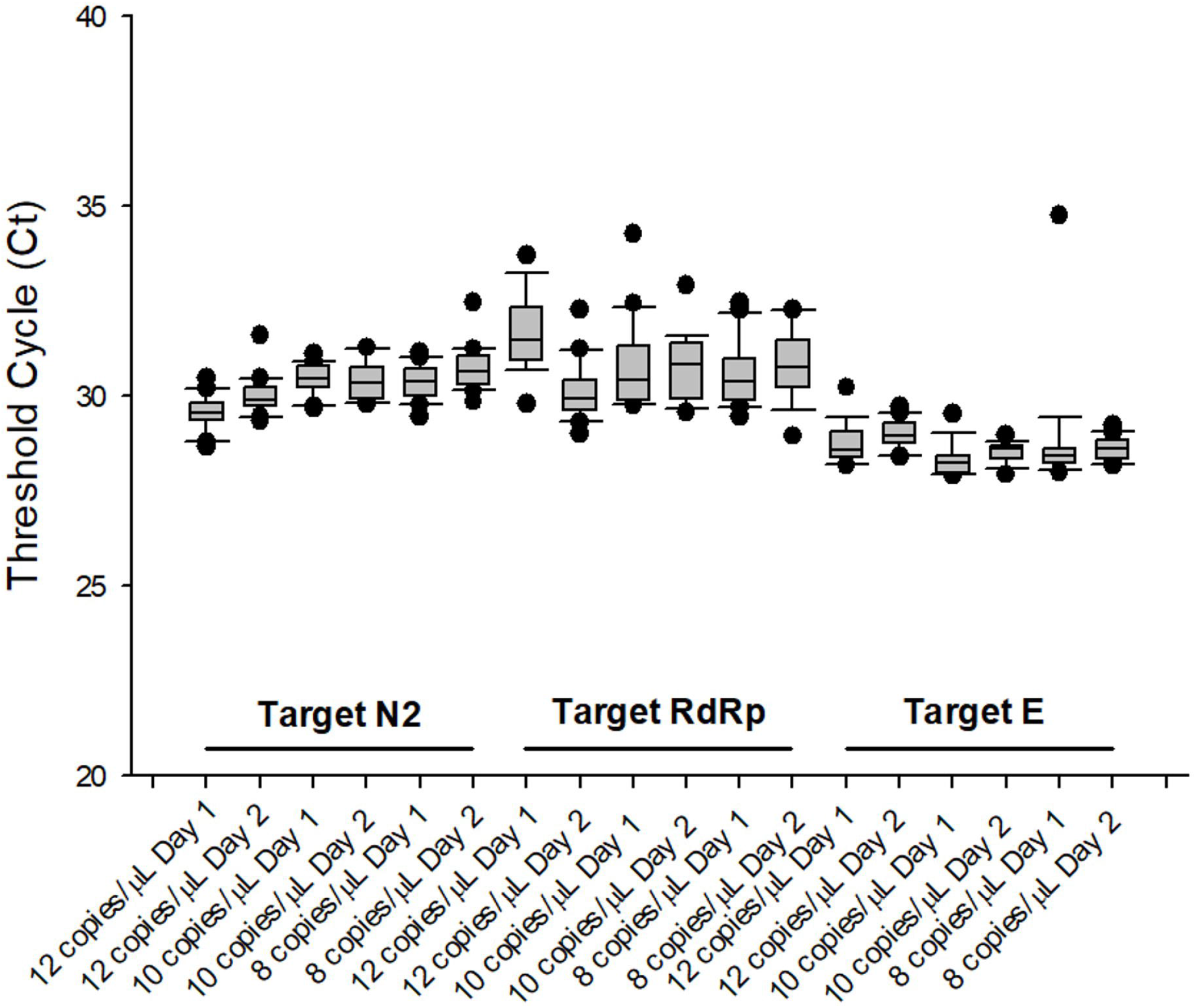
Reproducibility of HRM-RTqPCR assays targeting SARS-CoV-2 E, N and RdRp. SARS-CoV-2 templates were assayed in 40 technical replicates, at 12, 10 and 8 copies/µL.

### Clinical validation of HRM-RTqPCR assays

In order to assess the quality of the samples and RNA extraction, differentiating true- and false-negative results, the amplification of the internal control human RNAse P was monitored in all clinical specimens, to the TaqMan RT-qPCR and HRM-RTqPCR assays, in parallel to the detection of SARS-CoV-2 (Fig. 3A). Even considering some outliers, no significant difference was observed in Ct values between TaqMan and HRM assays (p= 0.634), showing the improved amplification of the RNAse P target in the HRM assay developed in this study. In addition, all the samples presented RNAse P Ct<35, except samples 54 (Ct 35.203 – serum) and 65 (Ct 36.440 – saliva) (Table S1). When the Ct values were compared between the type of clinical samples (swab, serum or saliva – Fig. 3B), significantly higher Ct values were observed to the serum samples, in comparison to swab or saliva. No significant difference was observed between swab and saliva samples (p= 1.000) (Fig. 3B), indicating a similar RNA quantity and recovery between both type of samples.

**Figure 3.**
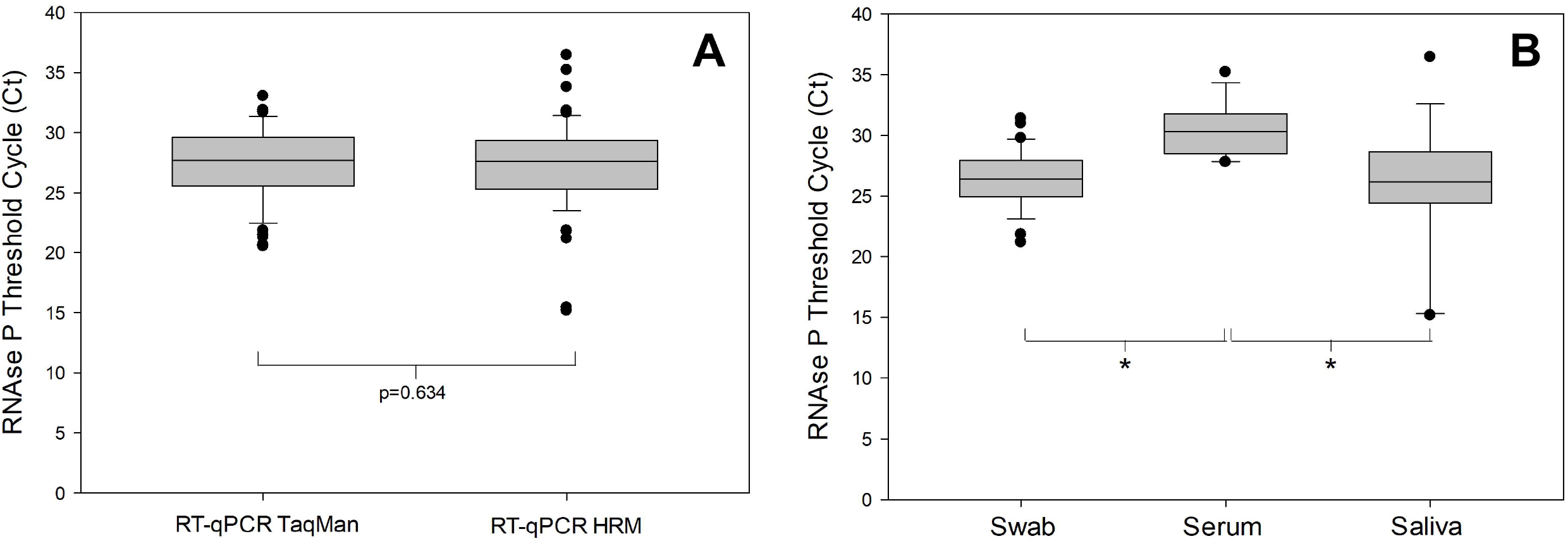
Box-Plot of RNAse P Ct values between RT-qPCR TaqMan and RT-qPCR HRM assays and between swab, serum and saliva samples. The Ct values from the amplification of the internal control human RNAse P was compared between TaqMan and HRM assays in all clinical specimens (A) or between swab, serum and saliva samples (B). The outliers are represented as the black circles. In A, p=0.634 (Mann-Whitney Rank Sum Test). In B, *p<0.05 (One-way ANOVA) and p=1.00 (Swab x Saliva - Mann-Whitney Rank Sum Test).

After assessing the quality of the samples through the human RNAse P internal control amplification, the clinical validation on the SARS-CoV-2 detection was performed with samples from 65 patients from the Rio de Janeiro and Ceará States in Brazil (42 swab, 12 serum and 11 saliva samples). The TaqMan assay targeting the N region of SARS-CoV-2 was considered the gold standard for the parameter’s evaluation. From the 65 samples, 51 were positive and 14 negative by the gold standard. The sensitivity of the HRM RTqPCR assays targeting N, RdRp and E were, respectively, 94.12, 98.04 and 92.16% (Table 3). The specificities were 100% to the 3 targets. The Positive Predictive Values were 100% to all targets, and the Negative Predictive Values were 82.35, 93.33 and 77.18% to the N, RdRp and E targets, respectively. In addition, the diagnostic accuracy was 95.38, 98.46 and 93.85%, respectively. To estimate the agreement between HRM x TaqMan assays, the Cohen kappa coefficient was calculated. It was observed the coefficients of 0.87, 0.96 and 0.84 to the N, RdRp and E targets, respectively. In general, the three HRM assays presented good clinical validation parameters, with elevated diagnostic accuracy, above 90%, in comparison to the TaqMan assay, including one positive amplification in serum samples (from 12) and 8 positive amplification in saliva samples (from 11). In conjunct, the HRM assay targeting RdRp was the one with the best clinical performance, with the most elevated sensitivity, specificity, PPV, NPV, accuracy and Cohen’s kappa coefficient levels.

**Table 3.** Clinical validation of RT-qPCR assays targeting SARS-CoV-2 E, N and RdRp. The parameters of clinical validation were calculated considering RT-qPCR with TaqMan assays as gold standard, using the OpenEpi (v. 3) software.

| Parameters | HRM RTqPCR assays |  |  |
| --- | --- | --- | --- |
|  | N target [CI95%] | RdRp target [CI95%] | E target [CI95%] |
| Samples number | 65 | 65 | 65 |
| Sensitivity (%) | 94.12 [84.08, 97.98] | 98.04 [89.7-99.65] | 92.16 [81.5-96.91] |
| Specificity (%) | 100 [78.47, 100] | 100 [78.47-100] | 100 [78.47-100] |
| PPV (%) | 100 [92.59, 100] | 100 [92.86-100] | 100 [92.44-100] |
| NPV (%) | 82.35 [58.97, 93.81] | 93.33 [70.18-98.81] | 77.18 [54.78-91] |
| Diagnostic accuracy (%) | 95.38 [87.29, 98.42] | 98.46 [91.79-99.73] | 93.85 [85.22-97.58] |
| Cohen's kappa coefficient | 0.87 [0.63-1.11] | 0.96 [0.71- 1.19] | 0.84 [0.60- 1.08] |

PPV: Positive predictive value, NPV: Negative predictive value, CI95%: 95% Confidence Intervals.

Finally, the pairwise comparison of Ct values between RT-qPCR TaqMan and HRM-RTqPCR assays to the N, RdRp and E targets in SARS-CoV-2 was performed using a Bland-Altman Plot (Fig. 4). The comparison was done to the TaqMan N x HRM N assays (Fig. 4A), TaqMan N x HRM RdRp assays (Fig. 4B) and TaqMan N x HRM E assays (Fig. 4C). The Bland-Altman Plot is a method to evaluate the agreement between two different assays, allowing the identification of any systematic difference between the measurements (i.e., fixed bias) or possible outliers. To the comparison between TaqMan and HRM to the N target for the 65 specimens analyzed, a bias of 0.56 and only three outliers (with Ct difference outside de mean±1.96 Standard Deviation) were observed (Fig. 4A). When the comparison was performed between the TaqMan N and HRM RdRp assays, a bias of −1.71 with only three outliers were observed (Fig. 4B). Lastly, when the comparison was between TaqMan N and HRM E assays, a bias of −0.89 with also three outliers were observed. In all comparisons, those parameters represent a very good agreement between the assays, which resulted in a performance of the HRM-RTqPCR similar to the gold standard TaqMan RTqPCR assay.

**Figure 4.**
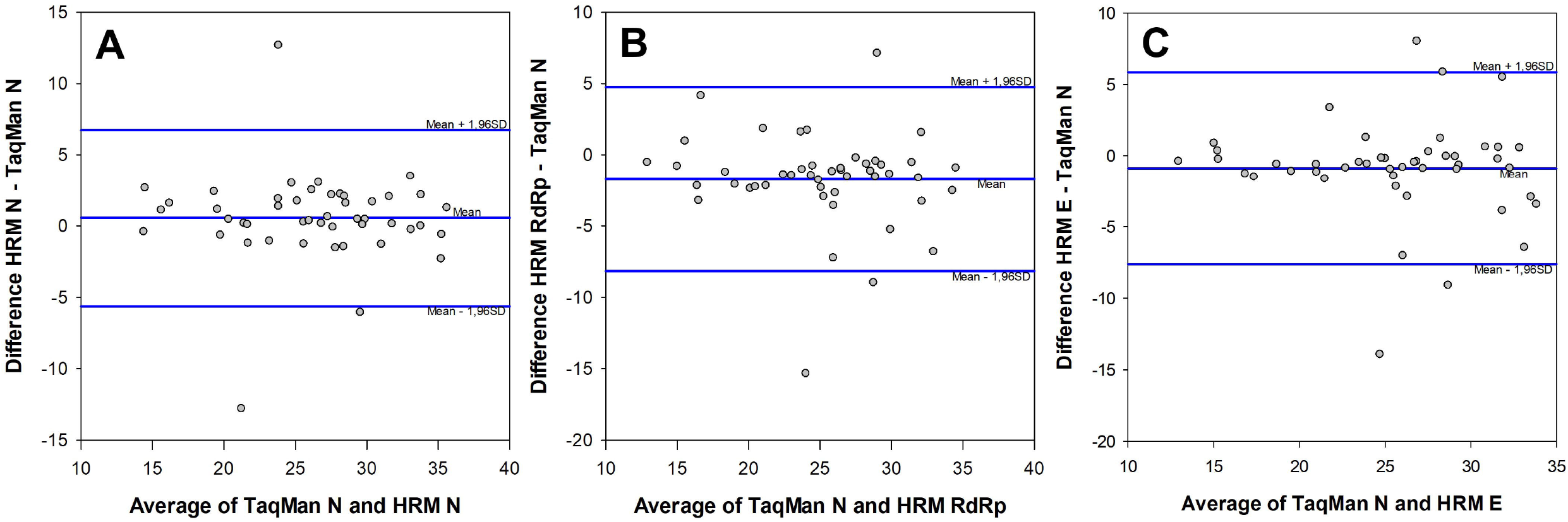
Bland-Altman Plots resulting from the pairwise comparison of Ct values between RT-qPCR TaqMan and HRM-RTqPCR assays targeting SARS-CoV-2 N, RdRp and E. A. TaqMan N x HRM N targets. B. TaqMan N x HRM RdRp targets. C. TaqMan N x HRM E targets. Level of agreement between two assays, also known as observed mean difference (blue line at center), is presented as a function of the average Ct values of the two assays. Upper and lower limits of agreement (Upper and lower blue lines, respectively) defined by mean Ct difference ± 1.96 Standard Deviation (SD). The Bias observed were 0.56, −1.71 and −0.89 to A, B and C, respectively.

## DISCUSSION

With the emergence of the Covid-19 pandemic, the world faced an unprecedented need for RT-qPCR-based molecular diagnostic tests, leading to a lack of kits and inputs, especially in developing countries. In March 2020, to increase the reach of Covid-19 molecular diagnostic, WHO produced a technical manual recommending seven in-house RT-qPCR assays, developed and validated by recognized research centers worldwide ^8^. This manual helped institutions of support, such as universities and small research groups, to also contribute to the Covid-19 frontline. However, with the high demand for molecular diagnostic tests, the costs for commercial kits and inputs were overrated, stimulating the development of alternative methods to detect SARS-CoV-2 in clinical specimens. Therefore, the main goal of this study was to develop a low-cost in-house HRM-RTqPCR assay, using alternative and more economical inputs, to increase the testing of patients suspicious for Covid-19, especially in restricted-budget laboratories.

HRM is a method developed in 2002 by the University of Utah (USA) and Idaho Technology (USA) for DNA sequence analysis ^9^. As the easiest DNA genotyping method, mutation screening and sequence matching, the number of studies reporting the use of HRM is continuous growth. The HRM cost-advantage results from the easiness of the method: beyond the PCR reagents, it is required only a saturating DNA dye, that costs much less than a fluorescent probe. Although genotyping corresponds to the main HRM application, the use of this methodology for the simultaneous molecular diagnostic and genotyping of pathogens, including the first coronavirus, is also promising ^10, 13-15^.

Based on the TaqMan in-house assays, we first evaluated all the primers sequences *in silico*, selecting three primers pairs targeting the N, RdRp and E regions of SARS-CoV-2 genome. As the main objective of this study was to develop HRM-RTqPCR assays to the molecular detection of the virus, regardless the mutations accumulated by the variants of concern, it was important to select the most stable genes to the diagnostic assays, to avoid the presence of insertions, deletions or SNPs that can lead to primer mismatches or differences in the shape of the HRM curves, used to certify the specificity of the RT-qPCR amplifications.

In this study, the analytical validation of HRM-qPCR assays was performed based on international guidelines ^16^, using synthetic sequences of SARS-CoV-2. Our first results showed a sharp peak for each SARS-CoV-2 target and human RNAse P at the derivative melting curves, with Tms differing at least 2.5°C between the targets. Although it was not explored in this study, the Tm differences open a possibility for multiplexing these assays, lowering even more the costs. However, in order to reach the best condition, it could be necessary to standardize the assays in multiplex, modifying the concentrations of primers and master mix reagents, as MgCl2 and dNTP, which is limited in the commercial HRM master mixes available.

The dynamic range of the HRM-RTqPCR assays observed to the 3 SARS-CoV-2 targets presented 5 logs and approximated LOD of 10 copies/µL, with PCR efficiencies between 88.3% and 109.7%. The shape of the normalized HRM curves was very similar between all positive patient samples and the synthetic templates, used as reference. In fact, the clinical specificity observed to those three HRM-RTqPCR assays were 100%, which corroborates the similar shape and solely peak of each HRM curve. If a mutation occurred in the N, RdRp and E regions amplified by the primers used, it would be possible to observe a difference in the normalized HRM curves, which would be detected by the HRM software and reported as a variation. In this study, by the 145 positive HRM-RTqPCR results, the stability of the N, RdRp and E regions in the SARS-CoV-2 genome was demonstrated.

One of the main concerns in HRM-based assays regards its repeatability and reproducibility. Following the Clinical and Laboratory Standards Institute (CLSI) document EP12-A2 for qualitative tests ^17^, we performed a replication experiment to evaluate precision of the three HRM-RTqPCR assays to SARS-CoV-2 using three low concentrations of the synthetic templates, around the estimated LOD. The parameters of precision were evaluated from the Ct values observed in each assay. The three assays presented a high precision, estimated by the small coefficient of variation observed, below 2%, except for the E target assay at 8 copies/µL (3.65%). Even for this target, the CV was below 15%, as recommended ^18^. To evaluate reproducibility, the three assays were performed in twenty technical replicates for each concentration around the LOD, by the same operator in two consecutive days, totalizing forty replicates. The Ct values were compared between day one and day two for each target, but no statistical difference was observed (Fig. 2), assuring the good reproducibility of the method.

The clinical validation of the HRM-RTqPCR assays was performed with 65 samples, mostly nasopharyngeal swabs, but also saliva and serum. The human RNAse P gene was used as internal control, for both TaqMan and HRM assays. No statistical difference was observed in the RNAse P Ct values between TaqMan and HRM assays, which suggest that the sensitivity for this target amplification was similar between the two methods. Nevertheless, when the Ct values were compared between the different type of samples in the HRM-RTqPCR assays, serum samples presented the higher RNAse P Ct values, significantly different from nasopharyngeal swabs and saliva. As expected, during the serum preparation, cells containing mRNA, including the RNAse P transcript, keep trapped at the clot. This probably result in a lower detection of this human target in serum samples. Regardless, previous studies already described detection of SARS-CoV-2 in serum ^19^, which could be associated with increase mortality risk in hospitalized COVID-19 patients ^20^.

As reported, the three HRM assays validated herein presented high sensitivity (from 92.16% to 98,04%) and specificity (all 100%). Regardless the kind of sample, a very good concordance could be observed with the TaqMan assay, except for three nasopharyngeal swab and one serum samples (Table S1). From those, only one sample was positive to TaqMan but negative to the three HRM assays. This sample presented Ct=37.02±0.07, the highest Ct observed to the TaqMan assays in this study. In addition, the other three samples also presented high Ct values to the TaqMan assay, higher than It is important to mention that most patients suspicious for Covid-19 that search the health systems are in the acute phase of the disease, in which a high viral load is observed and the molecular diagnostic is recommended ^21^. In addition, the Ct values variability among the the samples observed in this study can, perhaps, be justified due the fact that the samples analyzed were not collected during similar time intervals between the symptoms onset and collection. Noteworthy, the HRM-RTqPCR assays validated in this study presented elevated sensitivity to the detection of the SARS-CoV-2 genome even considering the samples presenting lower viral loads, observed through high Ct values in the gold standard assay.

Finally, the agreement between TaqMan and HRM assays was evaluated by the Bland-Altman plot analysis with the pairwise comparison of Ct values. The HRM assays to three targets in the virus were compared to the TaqMan assay targeting the N region. The three HRM assays present a high agreement with the gold standard, with biases very close to zero and only three outliers (out of ±1.96 Standard Deviation) in each comparison. It means that the Ct values are very similar performing the TaqMan or HRM assays, even for different targets in the SARS-CoV-2 genome. Considering that each region presents only one copy at the virus genome, and the PCR efficiencies were very similar among the targets and close to 100%, it is plausible to expect similar Ct values to all the targets and the TaqMan assay. Taken together, our results open a new venue of low-cost alternatives for the molecular diagnostic of Covid-19 in different kind of samples, with promising application to the SARS-Cov-2 genotyping and identification of the variants of concern, after the adaptation of the current assays to the S region in the viral genome. Of note, the estimated cost for the HRM-RTqPCR assay is significantly lower than a commercial kit for the molecular diagnostic of Covid-19 based on the TaqMan system, and much lower than a DNA sequencing to identify the SARS-CoV-2 variants, which could represent a considerable economy for the public health systems.

### Conclusion

The HRM-RTqPCR assays targeting the N, RdRp and E regions of the SARS-CoV-2 presented high sensitivity and specificity to the molecular diagnostic of Covid-19 from nasopharyngeal swabs, serum or saliva samples, being a low cost alternative to the TaqMan assays current available and a promising methodology to the development of new assays to the discrimination of the SARS-CoV-2 variants of concern.

## Data Availability

All relevant data is available in the manuscrit and supporting tables

## ACKNOWLEDGMENTS

Authors would like to thank to all the clinical staff involved in patient assistance and sample harvesting, as well as the technicians from the Technological Platforms from Fiocruz for the support.

## AUTHOR CONTRIBUTIONS

OM and LL are the guarantors of this work and, as such, had full access to all of the data in the study and take responsibility for the integrity of the data and accuracy of data analysis. OM conceived and designed the study. SVAL take the responsibility of write and submit the approved COVID-19 Biomarker Project, RK and MA admitted and followed-up part of the Covid-19 patients, obtained samples for the qPCR assays; OM Analyzed the oligonucleotides and designed the synthetic templates; WC, VDC, LMV VC) carried out the RNA extraction from nasopharyngeal swabs, serum and saliva samples; BF and NG carried out experiments related to standardization and validation of the HRM-RTqPCR assays; LMV, FM, VP and LL supervised analysis and interpretation of results and revised the manuscript. OM wrote the original draft and the revised manuscript. OM and LL gave financial support to the work. All authors approved the final version of the manuscript.

## SUPLEMENTARY TABLES

**Table S1.**
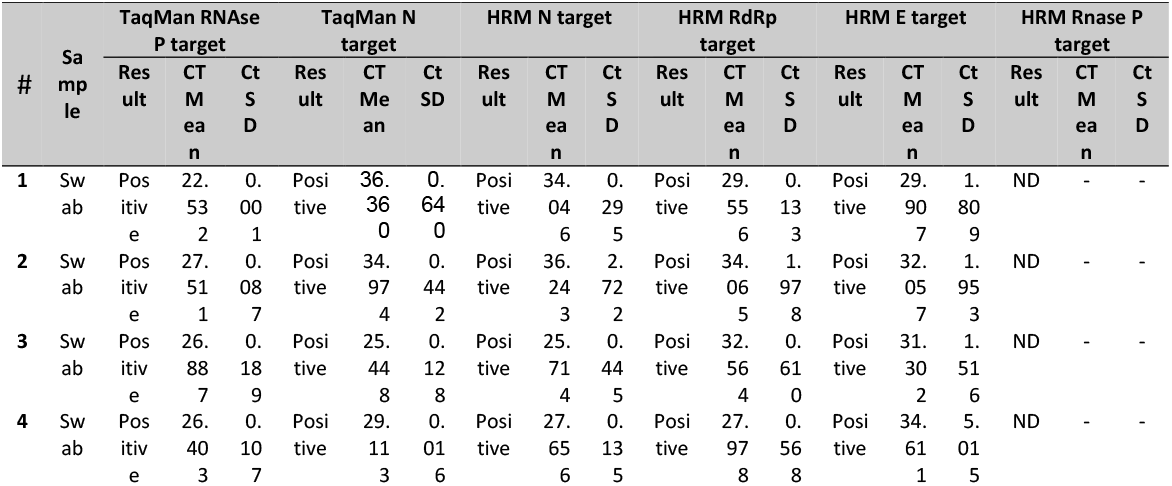
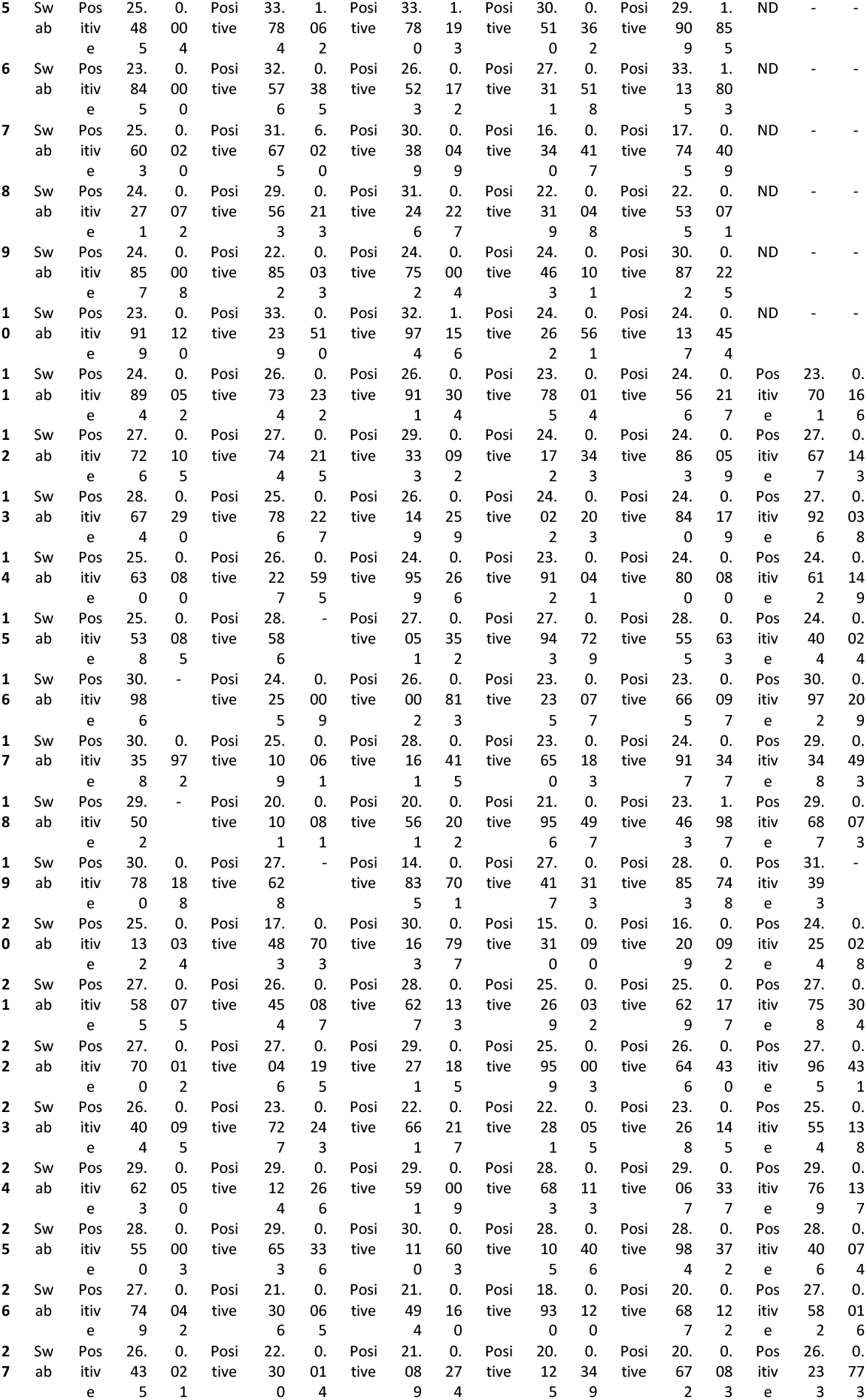
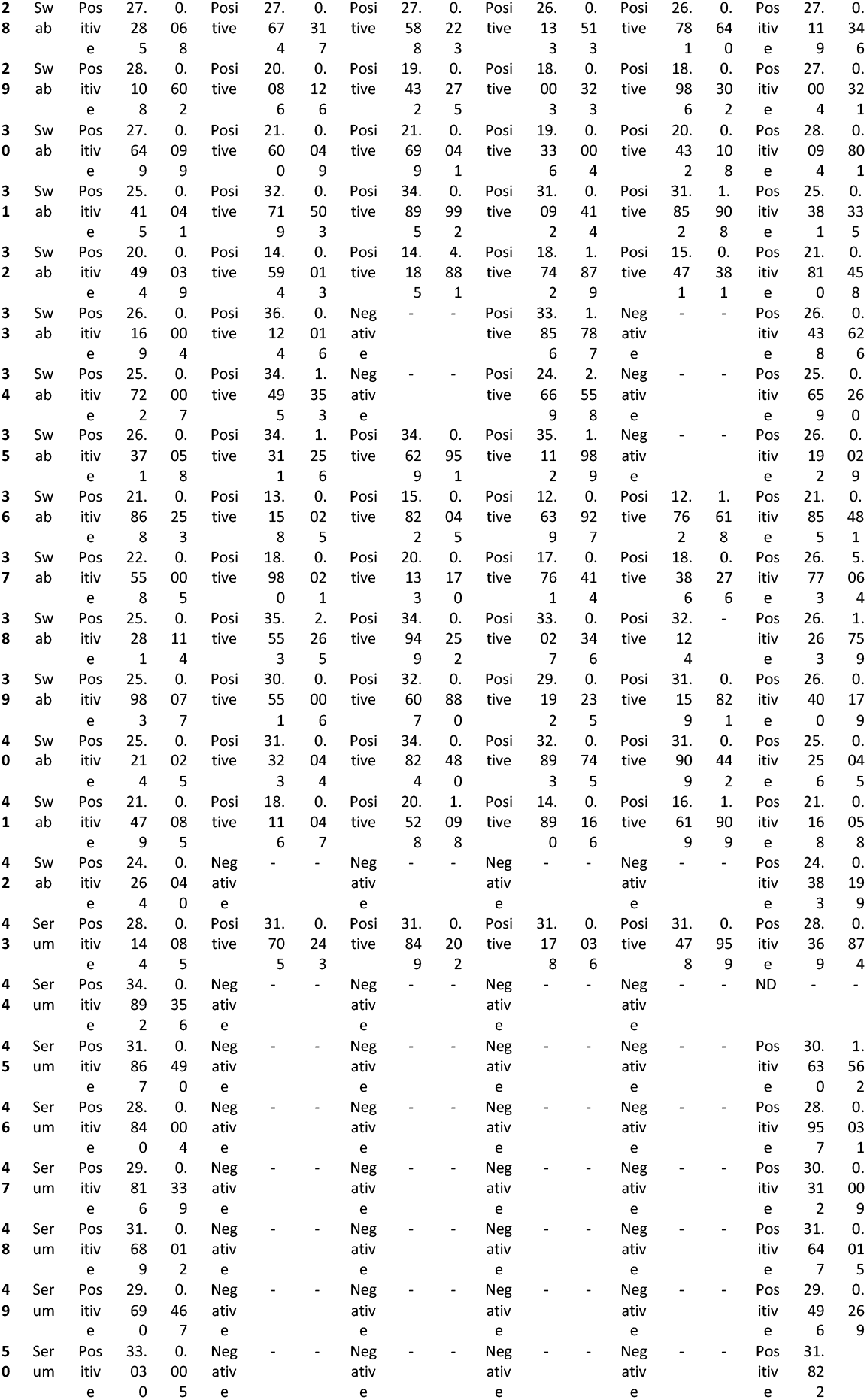
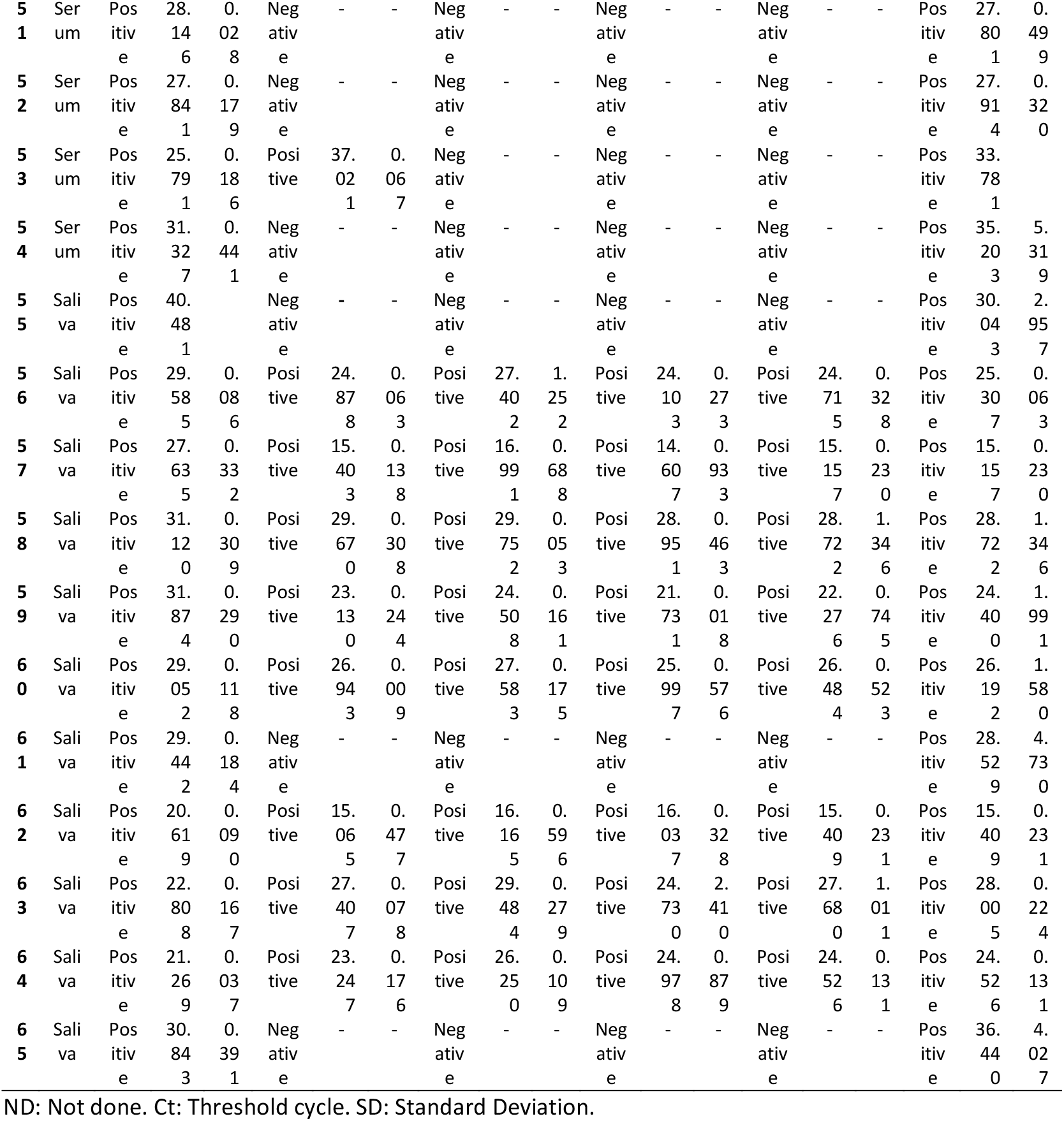
Validation of the HRM-RTqPCR assays for SARS-CoV-2 E, N and RdRp targets. Sixty-five RNA samples were extracted from nasopharyngeal swab, serum or saliva of patients suspicious for Covid-19. All samples were analyzed for RT-qPCR TaqMan assays targeting SARS-CoV-2 N2 and Human RNAseP targets, in parallel to the HRM-RTqPCR assays.

## REFERENCES

1. Machhi J, Herskovitz J, Senan AM, Dutta D, Nath B, Oleynikov MD, et al.: The Natural History, Pathobiology, and Clinical Manifestations of SARS-CoV-2 Infections. Journal of Neuroimmune Pharmacology 2020, 15 (3): 359–386.

2. Gorbalenya AE, Baker SC, Baric RS, de Groot RJ, Drosten C, Gulyaeva AA, et al.: The species Severe acute respiratory syndrome-related coronavirus: classifying 2019-nCoV and naming it SARS-CoV-2. Nature Microbiology 2020, 5 (4): 536–544.

3. WHO. COVID-19 Weekly Epidemiological Update (May 11th, 2021). World Health Organization 2021. Available at: https://www.who.int/publications/m/item/weekly-operational-update-on-covid-19. Accessed at 5-july-2021

4. Morbidity and Mortality Weekly Report (MMWR). Coronavirus disease 2019 in children – United States 2020, 69 (14):422–426.

5. Lu R, Zhao X, Li J, Niu P, Yang B, Wu H, Wang W, Song H, Huang B, Zhu N, Bi Y, Ma X, Zhan F, Wang L, Hu T, Zhou H, Hu Z, Zhou W, Zhao L, Chen J, Meng Y, Wang J, Lin Y, Yuan J, Xie Z, Ma J, Liu WJ, Wang 441 D, Xu W, Holmes EC, Gao GF, Wu G, Chen W, Shi W, Tan W: Genomic characterization and epidemiology of 2019 novel coronavirus: implications for virus origins and receptor binding. Lancet 2020, pii: S0140-6736(20)30251-8.

6. Wu F, Zhao S, Yu B, Chen YM, Wang W, Song ZG, Hu Y, Tao ZW, Tian JH, Pei YY, Yuan ML, Zhang YL, Dai FH, Liu Y, Wang QM, Zheng JJ, Xu L, Holmes EC, Zhang YZ: A new coronavirus associated with human respiratory disease in China. Nature 2020, 579: 265–269.

7. Corman VM, Landt O, Kaiser M, Molenkamp R, Meijer A, Chu DK, Bleicker T, Brünink S, Schneider J, Schmidt ML, Mulders DG, Haagmans BL, van der Veer B, van den Brink S, Wijsman L, Goderski G, Romette JL, Ellis J, Zambon M, Peiris M, Goossens H, Reusken C, Koopmans MP, Drosten C: Detection of 2019 novel coronavirus (2019-nCoV) by real-time RT-PCR. Euro Surveill 2020, 25.

8. WHO 2020. WHO in house protocols for COVID-19. World Health Organization. Available at: https://www.who.int/docs/default-source/coronaviruse/whoinhouseassays.pdf?sfvrsn=de3a76aa_2. Acessed at: 26-april-2020.

9. Reed GH, Kent JO, Wittwer CT: High-resolution DNA melting analysis for simple and efficient molecular diagnostics. Pharmacogenomics 2007, 8(6): 597–608.

10. Hewson KA, Browning GF, Devlin JM, Ignjatovic J, Noormohammadi AH: Application of high-resolution melt curve analysis for classification of infectious bronchitis viruses in field specimens. Aust Vet J 2010, 88(10):408–13.

11. Tong SYC, Giffard PM: Microbiological Applications of High-Resolution Melting Analysis. Journal of Clinical Microbiology 2012, 50(11): 3418–3421.

12. Hong YJ et al.: Detection of herpes simplex and varicella-zoster virus in clinical specimens by multiplex real-time PCR and melting curve analysis. BioMed Research International 2014, 2014: 261947.

13. Sun Y, Cheng Y, Lin P, Zhang H, Yi L, Tong M, Cao Z, Li S, Cheng S, Wang J: Simultaneous detection and differentiation of canine parvovirus and feline parvovirus by high resolution melting analysis. BMC Vet Res 2019, 15(1):141.

14. Hewson, K., Noormohammadi, A.H., Devlin, J.M. et al.: Rapid detection and non-subjective characterisation of infectious bronchitis virus isolates using high-resolution melt curve analysis and a mathematical model. Arch Virol 2009, 154: 649.

15. Chua, K. H. et al.: Development of High Resolution Melting Analysis for the Diagnosis of Human Malaria. Sci Rep 2015, 5: 15671.

16. Burd EM: Validation of laboratory-developed molecular assays for infectious diseases. Clin Microbiol Rev 2010, 23(3): 550–76.

17. CLSI/NCCLS 2008: User protocol for evaluation of qualitative test performance. Approved guideline, 2nd ed. CLSI document EP12-A2. Clinical and Laboratory Standards Institute 2008., Wayne, PA.

18. Food and Drug Administration 2001: FDA guidance for industry: bioanalytical method validation. Center for Drug Evaluation and Research, U.S. Department of Health and Human Services, Rockville, MD.

19. Kima JM, Kima HM, Leea EJ, Joa HJ, Yoona Y, Leea NJ, Sona J, Leea YJ, Kima MS, Leea YP, Chaea SJ, Parka KR, Choa SR, Parka S, Kima SJ, Wanga E, Wooa SH, Lima A, Parka SJ, Janga JH, Chunga YS, Chinb BS, Leec JS, Limd D, Hana MG, Yoo CK: Detection and Isolation of SARS-CoV-2 in Serum, Urine, and Stool Specimens of COVID-19 Patients from the Republic of Korea. Osong Public Health Res Perspect 2020, 11(3):112–117.

20. Rodríguez-Serrano DA, Roy-Vallejo E, Zurita Cruz ND, Martín Ramírez A, Rodríguez-García SC, Arevalillo-Fernández N, Galván-Román JM, Fontán García-Rodrigo L, Vega-Piris L, Chicot Llano M, Arribas Méndez D, González de Marcos B, Hernando Santos J, Sánchez Azofra A, Ávalos Pérez-Urria E, Rodriguez-Cortes P, Esparcia L, Marcos-Jimenez A, Sánchez-Alonso S, Llorente I, Soriano J, Suárez Fernández C, García-Vicuña R, Ancochea J, Sanz J, Muñoz-Calleja C, de la Cámara R, Canabal Berlanga A, González-Álvaro I, Cardeñoso L; REINMUN-COVID Group: Detection of SARS-CoV-2 RNA in serum is associated with increased mortality risk in hospitalized COVID-19 patients. Sci Rep 2021, 23, 11(1):13134.

21. Hanson KE, Caliendo AM, Arias CA, Hayden MK, Englund JA, Lee MJ, Loeb M, Patel R, El Alayli A, Altayar O, Patel P, Falck-Ytter Y, Lavergne V, Morgan RL, Murad MH, Sultan S, Bhimraj A, Mustafa RA: Infectious Diseases Society of America Guidelines on the Diagnosis of COVID-19: Molecular Diagnostic Testing. Infectious Diseases Society of America 2020, Version 2.0.0

